# Assessment of patient radiation dose in conventional lumbar spine radiography: A multicenter study in the Souss Massa region, Morocco

**DOI:** 10.64898/2026.03.24.26349174

**Authors:** Soudi Achraf, Menhour Youness

## Abstract

**Background:** Patient radiation exposure in diagnostic radiology is an important concern for radiation protection and patient safety. Monitoring radiation dose levels during radiographic examinations is essential to ensure compliance with diagnostic reference levels (DRLs) and to optimize radiological practices.

**Objective:** The aim of this study was to evaluate patient radiation dose during conventional lumbar spine radiography and compare the obtained values with diagnostic reference levels.

**Methods:** A descriptive cross-sectional multicenter study was conducted in four hospitals in the Sous Massa region, Morocco, between April and June 2017. Data were collected from 142 patients undergoing lumbar spine radiography examinations and from 20 radiology technicians. Exposure parameters including tube voltage, tube current, exposure time, focus-to-film distance, and field size were recorded. Entrance surface dose (ESD) was estimated using MICADO software, and dose area product (DAP) values were subsequently calculated. The 75^th^ percentile values were determined and compared with diagnostic reference levels.

**Results:** The regional 75^th^ percentile ESD values were 5.33 mGy for the anteroposterior projection and 7.38 mGy for the lateral projection. Corresponding DAP values were 1840.9 mGy.cm^2^ and 2783.65 mGy.cm^2^, respectively. All obtained values were below the diagnostic reference levels used for comparison. However, variations between hospitals were observed, likely due to differences in imaging protocols and equipment.

**Conclusion:** Radiation doses associated with lumbar spine radiography in the evaluated hospitals were within acceptable limits according to diagnostic reference levels. Continuous monitoring of patient radiation exposure and optimization of radiographic techniques remain essential to ensure effective radiation protection.

## INTRODUCTION

Radiological imaging plays a crucial role in modern medical diagnosis. However, diagnostic radiology is also one of the main sources of artificial radiation exposure to the population. According to the International Commission on Radiological Protection (ICRP), optimization of radiation dose in medical imaging is essential to minimize potential radiation risks while maintaining diagnostic image quality.

Diagnostic Reference Levels (DRLs) have been introducing as practical tools for radiation dose optimization in medical imaging. These reference levels help identify unusually high radiation doses and encourage healthcare institutions to optimize radiological procedures.

Lumbar spine radiography is among the most frequently performed radiographic examinations in clinical practice. Due to the relatively high radiation doses associated with this examination, monitoring patient exposure is particularly important.

Despite the increasing use of diagnostic reference levels worldwide, limited data are available regarding radiation exposure during conventional radiography in many developing regions. In Morocco, studies evaluating radiation doses during routine radiographic procedures remain scarce.

Therefore, this study aimed to assess patient radiation doses during conventional lumbar spine radiography in hospitals of the Souss Massa region and to compare the obtained values with diagnostic references levels.

## MATERIALS AND METHODS

### Study Design

This study was a descriptive cross-sectional multicenter investigation conducted between April and June 2017.

### Study setting

The study was carried out in four hospitals located in the Souss Massa region. These hospitals were selected based on their radiological activity and availability of conventional radiography equipment.

### Study Population

A total of 142 patients undergoing lumbar spine radiography were included in this study. Additionally, 20 radiology technicians participated in the data collection process.

### Data Collection

Data were collected using a standardized questionnaire and included:

- Patient demographic information
- Radiographic examination parameters
- Technical factors such as KV, mAs, and focus-to-film distance
- Field size and filtration parameters

### Dose Calculation

Entrance surface dose (ESD) was estimated using MICADO software, which calculates patient dose based on radiographic parameters. Dose Area Product (DAP) values were then calculated accordingly.

### Statistical Analysis

The 75^th^ percentile values of the measured doses were calculated to determine regional dose levels. These values were compared with diagnostic reference levels to evaluate compliance and dose optimization.

## RESULTS

A total of 142 patients undergoing conventional lumbar spine radiography were included in this study across four hospitals in the Souss Massa region.

### Patient Characteristics

The study population included adult patients undergoing lumbar spine radiography. The distribution of patients according to demographic characteristics is summarized in Table 1.

**Table 1:** Patient Characteristics.

| Variable |  | Value | Percentage (%) |
| --- | --- | --- | --- |
| Gender | Male | 74 | 52% |
|  | Female | 68 | 48% |
| Age | Moins de 19 ans | 46 | 32% |
|  | 40 à 59 ans | 55 | 39% |
|  | 20 à 39 ans | 32 | 23% |
|  | Moins de 19 ans | 9 | 6% |

Male patient accounted for 52% of the sample, while females represented 48%/the majority of patients included in the study were aged between 40 and 59 years, indicating that this age group was the most represented in the study population.

### Value of the entrance dose (De) and the dose-area product (DAP) for lumbar spine radiographic examinations

The table 2 demonstrates notable variability in entrance surface dose (De) and dose-area product (DAP) across the hospitals included in the study. Lateral (profile) projections consistently yielded higher dose values compared to anteroposterior (AP) projections. The highest recorded values were observed in Inzegane, with a mean DAP of 3343.51 mGy.cm^**2**^ and a 75^th^ percentile of 3394.97 mGy.cm^**2**^ for lateral projections. Similarly, the highest entrance dose (De) at the 75^th^ percentile reached 8.37 mGy in Inzegane for lateral views.

**Table 2:**
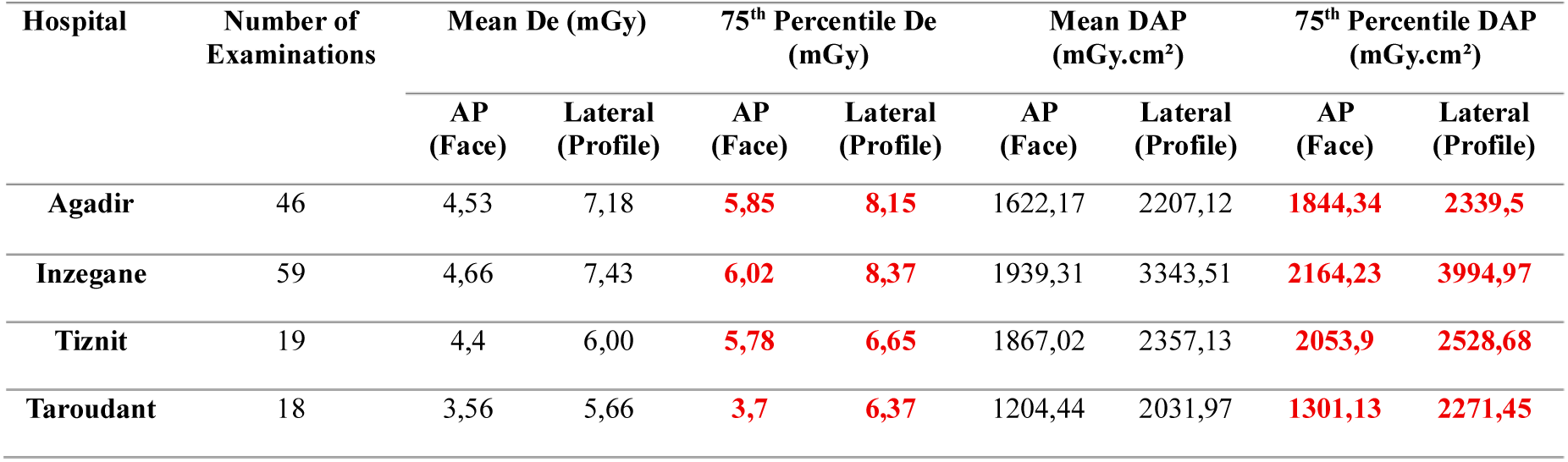
Entrance surface dose (De) and Dose-Area Product (DAP) values for lumbar spine radiographic examinations (anteroposterior and lateral projections), according to the hospitals included in the study.

**Table 3:**
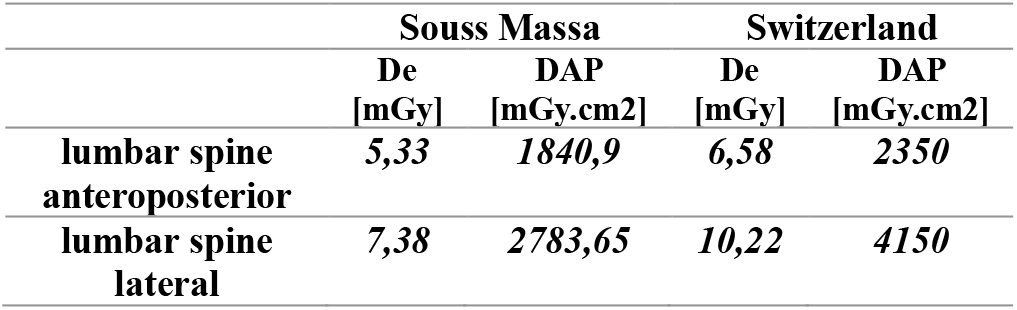
Comparison of entrance dose (De) and dose-area product (DAP) values in the Souss Massa region with those reported in Switzerland (Final report of the NRDRad project, Federal Office of Public Health FOPH, Consumer Protection Directorate, Radiation Protection Division).

|  | Souss Massa |  | Switzerland |  |
| --- | --- | --- | --- | --- |
|  | De<br>[mGy] | DAP<br>[mGy.cm2] | De<br>[mGy] | DAP<br>[mGy.cm2] |
| lumbar spine<br>anteroposterior | 5,33 | 1840,9 | 6,58 | 2350 |
| lumbar spine<br>lateral | 7,38 | 2783,65 | 10,22 | 4150 |

In contrast, the lowest values were reported in Taroudant, particularly for AP projections, with a mean De of 3.56 mGy and a mean DAP of 1204.44 mGy.cm^2^.

**Figure 1:**
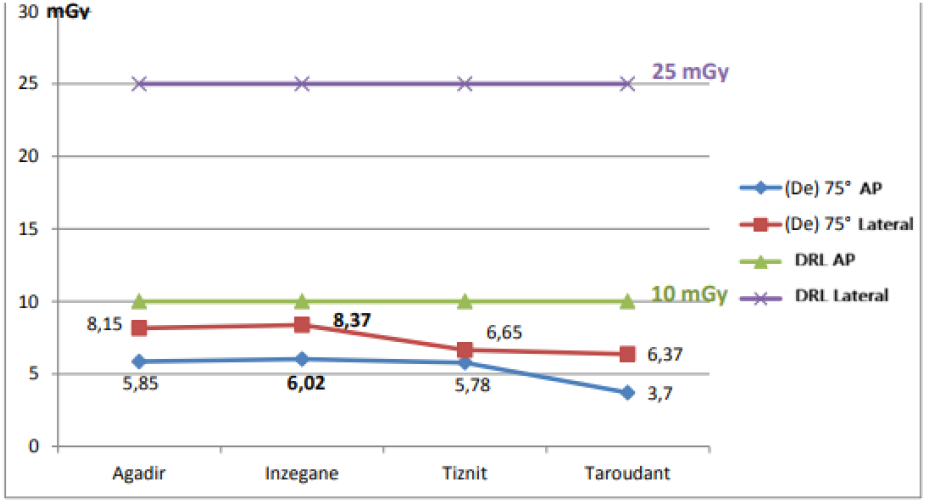
Distribution of 75^th^ percentile entrance dose (De) in mGy according to the hospitals and Diagnostic Reference Levels (DRL).

### Comparison of the values with Diagnostic Reference Levels (DRL)

The highest 75^th^ percentile of the entrance dose (De) for lumbar spine examinations was recorded in the radiology department of Inzegane, with values of 6.02 mGy for anteroposterior projection and 8.37 mGy for the lateral projection. Both values remain within the established Diagnostic Reference Levels (DRL).

### Distribution of the 75^th^ percentile dose-area product (DAP) according to hospitals (mGy.cm^2^)

**Figure 2:**
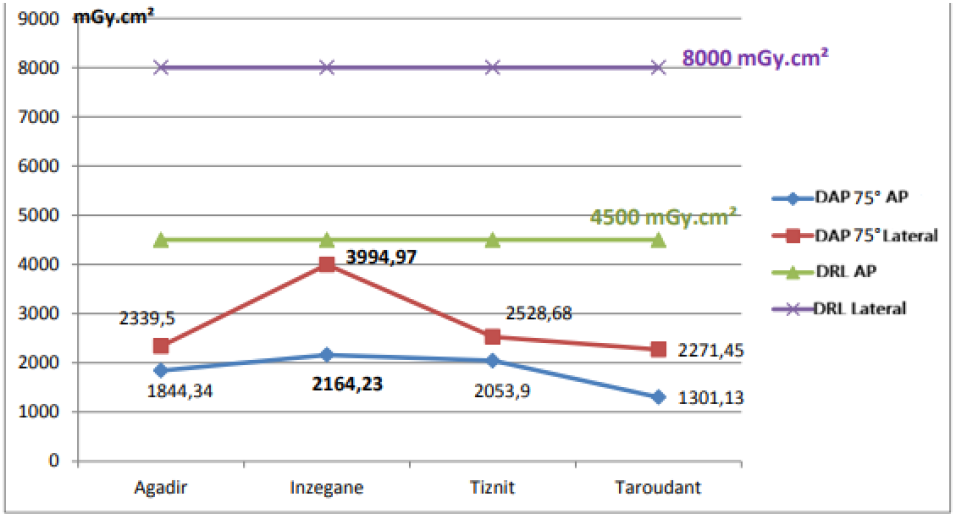
Distribution of the 75^th^ percentile dose-area product (DAP) according to hospitals and Diagnostic Reference Levels (DRL).

The highest 75^th^ percentile dose-area product (DAP) for lumbar spine examinations was recorded in the radiology department of Inzegane, with values of 2164.23 mGy.cm^2^ for the anteroposterior projection and 3994.97 mGy.cm^2^ for the lateral projection. Both values remain within the established Diagnostic Reference Levels (DRL).

### Distribution of the 75^th^ percentile entrance dose (De) for lumbar spine anteroposterior projections, calculated among patients in the Souss Massa region

**Figure 3:**
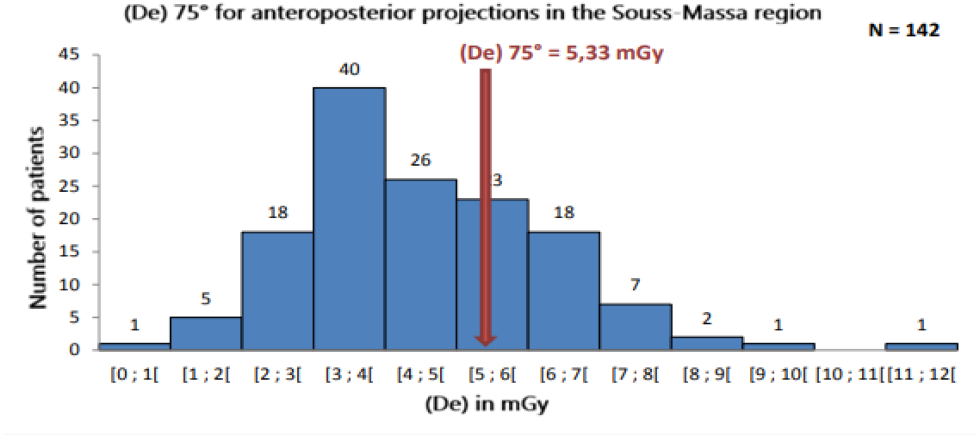
Distribution of the 75^th^ percentile entrance dose (De) for lumbar spine anteroposterior projections in the souss Massa region.

The 75^th^ percentile of the entrance dose (De) for lumbar spine anteroposterior projections in the Souss Massa region was 5.33 mGy.

### Distribution of the 75^th^ percentile entrance dose (De) for lumbar spine lateral projections, calculated among patients in the Souss massa region

**Figure 4:**
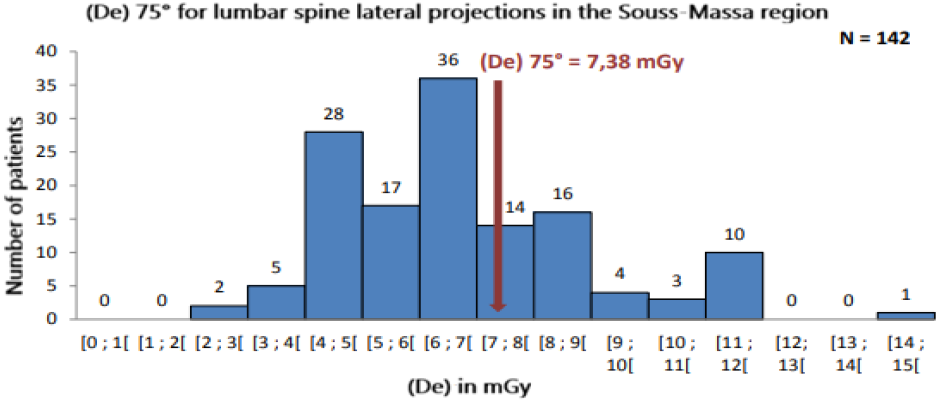
Distribution of the 75^th^ percentile entrance dose (De) for lumbar spine lateral projections in the souss Massa region.

The 75^th^ percentile of the entrance dose (De) for lumbar spine lateral projections in the Souss Massa region was 7.38 mGy.

### Distribution of the 75^th^ percentile dose-area product (DAP) for lumbar spine anteroposterior projections, calculated among patients in the Souss Massa region

**Figure 5:**
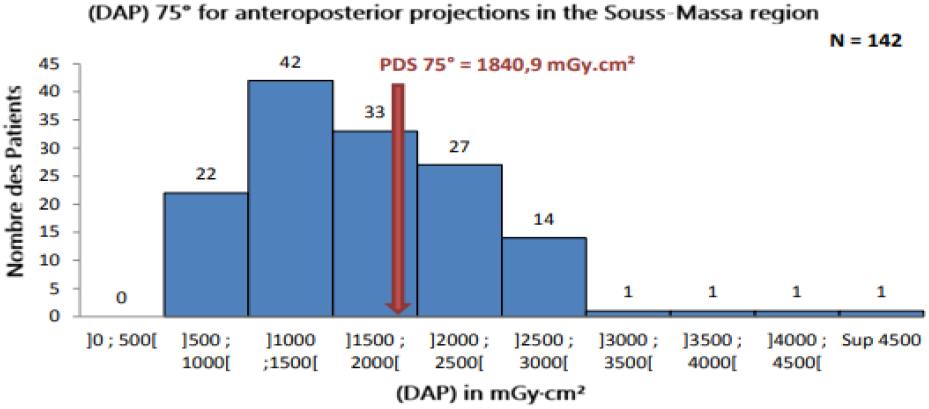
Distribution of the 75^th^ percentile (DAP) for lumbar spine anteroposterior projections in the souss Massa region.

The 75^th^ percentile of the dose-area product (DAP) for lumbar spine anteroposterior projections in the Sous Massa region was 1840.9 mGy.cm^2^.

### The distribution of the 75^th^ percentile dose-area product (DAP) for lumbar spine lateral projection, calculated among patients in the Souss Massa region

**Figure 6:**
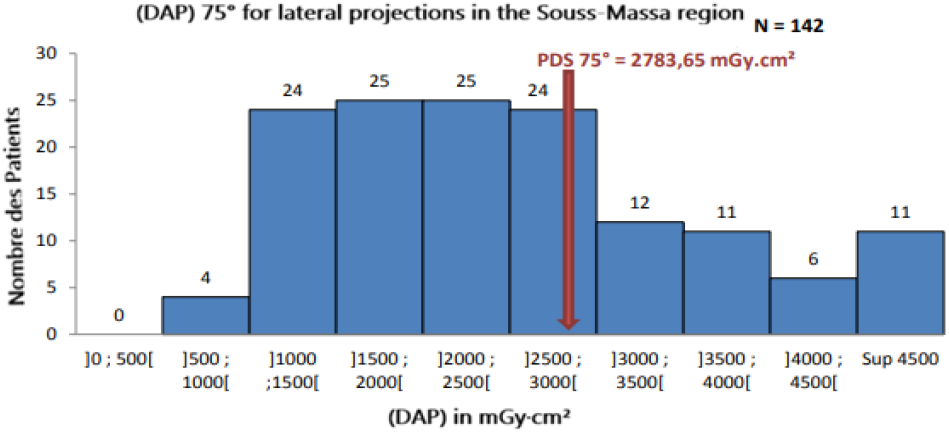
Distribution of the 75^th^ percentile (DAP) for lumbar spine lateral projections in the souss Massa region.

The 75^th^ percentile of the dose-area product (DAP) for lumbar spine lateral projections in the Souss Massa region was 2783.65 mGy.cm^2^.

## DISCUSSION

### A. Participant Characteristics

A total of 20 radiology technicians participated in this study out of 28 eligible professionals working across the radiology departements of fout hospitals in the Souss Massa region, resulting in a participation rate of 71%.

Among the participants, 75% was female, indicating that female staff are equally engaged in radiology services compared to their male counterparts, with a ratio of three women to one man.

The majority of participants (75%) has less the workforce is relatively young. In addition, 85% of the professionals reported having no knowledge of diagnostic reference levels (DRL), and all participants confirmed the absence of any guidelines or protocols related to dose optimization within their departements.

It was also observed that none of the professionals used fluoroscopy, and all examinations were performed with a focus-to-film distance (FFD) of 100 cm. Only the six technicians of radiology from

Inzegane reported that their departement had undergone a periodic evaluation in March 2017.

### B. Patient Parameters

Based on data collection forms, 142 patients were included in the study, with 52% male and 48% female.

In terms of age distribution, the majority of patients were within the 40-59 years age group (39%).

Radiology technicians adjusted examination parameters (KV, mAs, FFD, etc.) according to patient age, morphology, clinical condition, and examination conditions.

It was also noted that most professionals performed between 0 and 4 examination per day.

### C. Analysis of Calculated Doses

The collection data enabled the calculation of the 75^th^ percentile entrance surface dose (De) and dose area-product (DAP) for both the Souss Massa region and each participating departement. These values were compared with diagnostic reference levels (DRL) to assess dose optimization and compliance with radiation protection standards.

#### 1) Souss Massa region

The 75^th^ pecentile (De) values complied with DRL established by IRSN, with:

- **5.33** mGy for the anteroposterior projection (53.3% of DRL = 10 mGy), representing a reduction of 46.7%.
- **7.38** mGy for the lateral projection (29.52% of DRL = 25 mGy), representing a reduction of 70.48%.

Similarly, (DAP) values were also below DRL:

- **1840.9** mGy.cm^2^ for AP (40.9% of DRL = 4500 mGy.cm^2^), representing a reduction of 59.9%.
- **2783.65** mGy.cm^2^ for lateral (34.8% of DRL = 8000 mGy.cm^2^), representing a reduction of 65.2%.

#### 2) Results by Departement

All deppartements demonstrated compliance with DRL.

- **Agadir hospital (46 patients):** De: **5.85** mGy (AP), **8.15** mGy (LAT) DAP: **1844.34** mGy.cm^2^ (AP), **2339.5** mGy.cm^2^ (LAT)
- **Inzegane hospital (59 patients):** De: **6.02** mGy (AP), **8.37** mGy (LAT) DAP: **2164.23** mGy.cm^2^ (AP), **3994.97** mGy.cm^2^ (LAT)
- **Tiznit hospital (19 patients):** De: **5.78** mGy (AP), **6.65** mGy (LAT) DAP: **2053.9** mGy.cm^2^ (AP), **2528.68** mGy.cm^2^ (LAT)
- **Taroudant hospital (18 patients):** De: **3.7** mGy (AP), **6.37** mGy (LAT) DAP: **1301.13** mGy.cm^2^ (AP), **2271.45** mGy.cm^2^ (LAT)

#### 3) Departement Ranking

Based on dose optimization:

***De ranking***: Inzegane > Agadir > Tiznit > Taroudant

***DAP ranking***:Inzegane > Tiznit > Agadir > Taroudant

#### 4) Factor Influencing Dose Variation

The observed difference between departments can be explained by several factor:

- X-ray tube characteristics (voltage capacity, inherent and additional filtration): increased filtration reduces De and DAP.
- Detector performance and digital systems: more sensitive detectors allow reduced exposure parameters.
- Radiographic protocols:
  ➢ Increasing KV reduces De (if detector dose is constat)
  ➢ Increasing distance reduces De
  ➢ Increasing mAs increases dose and DAP

It should be noted that DAP is directly proportional to the irradiation field size.

#### 5) Comparison with International Data

Comparaison with a Swiss study conducted by the Federal Office of Public Health (2012) showed similar results, with both studies complying with DRL recommended by IRSN.

## CONCLUSION

This study aimed to update radiation doses delivered to patients in the Souss Massa region in diagnostic radiology, particularly in conventional lumbar spine radiography. The 75^th^ percentile entrance surface dose (De) and dose-area product (DAP) were determined for both the region and each participating departement and compared with diagnostic reference levels (DRL). The results showed that all values were below the recommended DRL, indicating good dose optimization and acceptable radiological practices. A reduction greater than 50% in both De and DAP was observed across all hospitals. These finding confirm satisfactory performance in radiation protection within the region. However, continuous monitoring of radiation doses remains essential to ensure sustained compliance with DRL. Periodic evaluation of radiological practice should be implemented to follow the evolution of techniques. In addition, staff training in dose calculation and DRL is necessary to improve knowledge and pratice. The involvement of a medical physicist or dosimetrist is strongly recommended to enhance radiation safety.

## Data Availability

All data produced in the present study are available upon reasonable request to the authors.

## DECLARATIONS

## ACKNOWLEDGEMENTS

The authors gratefully acknowledge the support of the regional health authorities and the heads of radiology departments in Agadir, Inzegane, Tiznit, and Taroudant for providing the necessary environment to conduct this multicenter research. Special appreciation is also extended to the patients and the 20 radiology technicians who participated in the data collection process.

## DATA AVAILABILITY

The data supporting the finding of this study (dosimetric parameters and MICADO software caluculation) are available from the corresponding author upon reasonable request.

## BIBLIOGRAPHY

Pages, J., & van Loon, R. (1998). The European protocol on dosimetry in mammography: Applicability and results in Belgium. Radiation Protection Dosimetry, 80(1-3), 191–193. 10.1093/oxfordjournals.rpd.a032503

Vaillant, L. (2024). Le principe de limitation des doses et la tolérabilité du risque radiologique. Radioprotection 10.1051/radiopro/2024026

International Commission on Radiological Protection (ICRP). (2007). The 2007 recommendations of the International Commission on Radiological Protection. Annals of the ICRP, 37(2–4), 1–332. 10.1016/j.icrp.2008.08.001

Bourguignon, M., et al. (2025). Les niveaux de référence diagnostiques en imagerie médicale: des exigences pour l’optimisation de la radioprotection des patients. Radioprotection. 10.1051/radiopro/2025043

Rehel, J.-L., & Roch, P. (2003). Niveaux de référence diagnostiques: spécificités de la démarche française en radiologie. Radioprotection, 38(2), 187–200. 10.1051/radiopro:2003008

Roch, P., & Aubert, B. (2013). French diagnostic reference levels in diagnostic radiology, computed tomography and nuclear medicine: 2004–2008 review. Radiation Protection Dosimetry, 154(1), 52–75. 10.1093/rpd/ncs152

Maccia, C. (1990). La dose reçue par les patients au cours des examens de radiodiagnostic et son optimisation. Radioprotection, 25(2), 135–154. 10.1051/radiopro:1990027

Akanni, D. W. M. M., Savi de Tové, K.-M., Damien, B. G., Kiki, S. M., Adjadohoun, S. B. M. G., Yekpe-Ahouansou, P., & N’zi, K. P. (2021). Connaissances en radioprotection des travailleurs exposés aux rayonnements ionisants en milieu médical en Afrique francophone sub-saharienne. Radioprotection, 56(1), 43– 48. 10.1051/radiopro/2020067

Sanou, A., Nikiéma, Z., & Traoré, O. (2010). Application des règles de protection contre les rayons X dans les services de radiologie de Ouagadougou. Radiation Protection Dosimetry, 139(1–3), 420–423. 10.1016/j.radi.2010.02.025

Office fédéral de la santé publique (OFSP). (2012). Niveaux de référence diagnostiques en radiologie par projection (rapport final). https://www.bag.admin.ch/dam/fr/sd-web/Slb6tKFWOVQu/schlussbericht-drw-rad.pdf. Consulted (20/06/2017 at 00h41).

Institut de Radioprotection et de Sûreté Nucléaire (IRSN). (2011). Occupational exposure to ionizing radiation in France: 2010 report (Report No. DRPH/DIR/2011-19). https://recherche-expertise.asnr.fr/sites/default/files/documents/expertise/rapports_expertise/IRSN_bilan_annuel_travailleurs_2010.pdf. Consulted (22/04/2017 at 17h41).

Mbo Amvene, J., Daoba, J., Soumayah, B., Kouong, J., Kueté Fomekong, U., & Nko’o Amvene, S. (2017). Evaluation de la Dose d’Entrée des Rayons X aux Patients lors de la Radiographie du Thorax en Pédiatrie. HEALTH SCIENCES AND DISEASE, 18(1). 10.5281/hsd.v18i1.768

Akanni, D. W. M. M., Fachinan, O. H., Adjadohoun, S. B. M. G., Zamba, A. K. J. J., Kiki, S. M., & Savi de Tové, K.-M. (2025). Radiographie du thorax en pédiatrie: justification et doses d’entrée à Parakou en 2021. Radioprotection, 60(3), 262–267. 10.1051/radiopro/2025006

Adambounou, K., Amoussou, K., Katassou, K., & Sedo, K. (2022). Dosimétrie des patients en radiologie conventionnelle au Togo. Médecine Nucléaire, 46, 98. 10.1016/j.mednuc.2022.01.124

